# Trends in hepatitis C virus seroprevalence and associated risk factors among men who have sex with men in Montréal: results from three cross-sectional studies (2005, 2009, 2018)

**DOI:** 10.1101/2020.01.27.20018994

**Authors:** Charlotte Laniece Delaunay, Joseph Cox, Marina B. Klein, Gilles Lambert, Daniel Grace, Nathan Lachowsky, Mathieu Maheu-Giroux

## Abstract

**Introduction:** To eliminate the hepatitis C virus (HCV) by 2030, Canada must adopt a micro-elimination approach targeting priority populations, including gay, bisexual, and other men who have sex with men (MSM). HCV prevalence and risk factors among MSM populations are context-dependent, and accurately describing these indicators at the local level is essential if we want to design appropriate, targeted prevention and treatment interventions. We aimed first to estimate and investigate temporal trends in HCV seroprevalence between 2005-2018 among Montréal MSM, and then to identify the socio-economic, behavioural, and biological factors associated with HCV exposure among this population.

**Methods:** We used data from three bio-behavioural cross-sectional surveys conducted among Montréal MSM in 2005 (n=1,795), 2009 (n=1,258), and 2018 (n=1,086). To ensure comparability of seroprevalence estimates across time, we standardized the 2005 and 2009 time-location samples to the 2018 respondent-driven sample. Time trends overall and stratified by HIV status, history of injection drug use (IDU), and age were examined. Modified Poisson regression analyses with generalized estimating equations were used to identify factors associated with HCV seropositivity pooling all surveys. We used multiple imputation by chained equations for all missing values.

**Results:** Standardized HCV seroprevalence among all MSM remained stable from 7% (95% confidence interval (CI): 3-10%) in 2005, to 8% (95%CI: 1-9%) in 2009, and 8% (95%CI: 4-11%) in 2018. This apparent stability hides diverging temporal trends in seroprevalence between age groups, with a decrease among MSM <30 years old, and an increase among MSM aged ≥45 years. History of IDU was the strongest predictor for HCV seropositivity (adjusted prevalence ratio: 8.0; 95%CI: 5.5-11.5), and no association was found between HCV seroprevalence and the sexual risk factors studied (condomless anal sex with men of serodiscordant/unknown HIV status, number of sexual partners, and group sex), nor with biological markers of syphilis.

**Conclusions:** HCV seroprevalence remained stable among Montréal MSM between 2005-2018. Unlike other settings where HCV infection was strongly associated with sexual risk factors among MSM subgroups, IDU was the preeminent risk factor for HCV seropositivity. Understanding the intersection of IDU contexts, practices, and populations is essential to prevent HCV transmission among MSM.

## Introduction

Canada endorsed the *World Health Organization*’s (WHO) global prevention and treatment service coverage targets to eliminate the hepatitis C virus (HCV) as a public health threat by 2030 [1]. With approximately 250,000 individuals chronically infected with HCV [2], Canada is not on track for HCV elimination [3]. The micro-elimination approach aims to develop tailored interventions to achieve the WHO’s targets among priority populations disproportionately affected by HCV [4, 5]. Due to exposures to HCV-contaminated blood and blood products before the introduction of HCV screening in 1990, HCV prevalence is high among individuals born between 1945-1975 in Canada [6]. Immigrants originating from HCV endemic countries may also be at increased risk of being chronically infected with HCV [7]. In the absence of behavioural risk factors such as sharing drug injection equipment, the risk of transmitting HCV is low among these groups. Contrarily, people who inject or use drugs, Indigenous peoples, people with experience in the prison system, and gay, bisexual, and other men who have sex with men (MSM) may be at high risk of acquiring and/or transmitting HCV [8].

Due to shared routes of transmission, HCV-HIV co-infection is common among individuals with similar vulnerabilities or risk behaviours. Since the early 2000s, HCV epidemics have been reported among MSM living with HIV in industrialized countries [9]. Global HCV incidence has more than doubled in this population, from 2.6/1,000 person-years before 2000 to 8.1/1,000 person-years since 2010 [10], with high reinfection rates [9]. Because HCV can be cleared, anti-HCV antibody detection in the blood –or seropositivity– is a marker of lifetime exposure to the virus, while detection of HCV RNA indicates an active infection. Global HCV seroprevalence among MSM living with HIV was estimated at 6.4% from 2002-2015 [11]. Comparatively, the burden of HCV has remained stable among HIV-negative MSM, with estimates of HCV seroprevalence ranging from 0.0-3.4% [12-16], and a global incidence of 0.4/1,000 person-years [17]. Nevertheless, recent cases of acute HCV infection have been reported among HIV-negative MSM without a history of injection drug use (IDU) in Europe [18], the United States [19], and Vancouver, British Columbia (Canada) [20]. These data may mask wide variations across geographical settings, highlighting the importance of local contexts and related risks.

In industrialized countries, most HCV infections have been attributed to the use of unsterile needles and drug paraphernalia, and receipt of blood or blood products before the introduction of HCV screening [21]. In parallel, evidence from Europe, the United States, and Vancouver shows that sexual transmission can account for a substantial proportion of incident HCV infections among MSM [9, 20, 22-24]. Socio-economic factors such as education and income can also influence the risk of acquiring HCV in this population [22, 25-27]. The impact of socio-economic factors and injecting and sexual behaviours on HCV transmission among MSM subgroups is context-dependent and, to date, there is little evidence of sexual transmission of HCV among MSM in Canada [15, 16, 20, 28, 29]. It is estimated that between 312,681-426,384 MSM live in Canada, representing approximately 2.6% of the adult male population [30]. The relatively large size of this key population, the variability in HCV incidence and prevalence among MSM populations, as well as the diversity of potential routes of transmission reflect the importance of using locally-valid epidemiological evidence to inform tailored interventions targeting micro-elimination.

Scarce evidence exists on HCV prevalence and risk factors among MSM subgroups in Canadian cities. In Vancouver, British Columbia, HCV seroprevalence among all MSM was estimated at 4.9% in a cross-sectional study conducted in 2008-2009 [15]. In this city, 16.8% of MSM living with HIV tested HCV seropositive, against 10.4% in a similar study conducted in Toronto, Ontario in 2010-2012 [16]. Temporal trends in HCV seroprevalence among MSM have not been assessed in Canada, however, available cross-sectional data suggest potential within-country variability in seroprevalence. In this study, we aimed to 1) investigate temporal trends in HCV seroprevalence among MSM living in Montréal from 2005-2018, overall and stratified by HIV status, IDU status, and age group; and 2) identify the social, behavioural, and biological factors associated with HCV exposure among this heterogeneous population.

## Methods

### Data sources and harmonization

Three cross-sectional surveys of HIV and other sexually transmitted and blood-borne infections were conducted among Montréal MSM in 2005, 2009, and 2018: Argus 1, Argus 2, and the first wave of the Engage cohort study, respectively. These surveys have been previously described [31, 32]. Briefly, they combined HIV, HCV, and syphilis surveillance and behavioural monitoring through self-administered questionnaires. Time-location sampling was used to recruit 1,957 MSM in 2005 and 1,873 in 2009. In 2018, a community-based sample of 1,179 MSM was recruited using respondent-driven sampling (RDS), a link-tracing, adaptive sampling method that uses information about the social networks of participants to estimate their probability of being recruited and to adjust for the biases associated with over/under-sampling of certain groups [33]. Eligibility criteria varied slightly across the three surveys (Additional file 1).

To compare the three surveys, we first harmonized the eligibility criteria across datasets. Our analyses were restricted to cis-gender men aged ≥18 years who reported sexual activity with a man in the past six months (P6M) and resided on the island of Montréal, Québec (Canada). The sensitivity and specificity of the assays used for anti-HCV antibody detection in the three surveys were comparable (sensitivity: 100% in 2005, 2009, and 2018; specificity: 99.95% in 2005 and 2009, and 99.69% in 2018 [31, 32, 34]).

### Temporal trends in HCV seroprevalence

We estimated HCV seroprevalence in 2005, 2009, and 2018, among all MSM, stratified by HIV status, lifetime history of IDU, and age group, and among HIV-negative MSM without a history of IDU. To ensure comparability of the samples across time, direct standardization was performed. The RDS-weighted 2018 survey was used as the standard and the other surveys were adjusted to yield the same distribution with regards to age, sexual orientation, annual income, and first language. This was achieved by first fitting multivariable logistic regression models with HCV seropositivity as the outcome (defined as a positive anti-HCV antibody test) to the 2005 and 2009 data, separately. Age was modeled using a restricted cubic spline [35], and we investigated the presence of multiplicative statistical interaction between each pair of covariates. For the stratified analyses, the same model was fitted separately among each subgroup of interest. Using these models, we then predicted the individual probability of testing HCV seropositive using the covariate patterns observed in the 2018 standard sample, among all MSM and by subgroup. Finally, we computed the weighted mean of these individual probabilities using Volz-Heckathorn weights [36] to estimate standardized HCV seroprevalence among the different groups at all time points. We obtained 95% confidence intervals (CIs) using cluster-level block bootstrap, with clusters corresponding to the recruitment site in 2005 (n=40 sites) and 2009 (n=39 sites) and to the initial seed of the recruitment chain in 2018 (n=27 seeds). We performed complete case analyses as only 5% of observations had any missing value for the variables used in these estimations [37].

### Social, behavioural, and biological factors associated with HCV seropositivity

Pooling data from the three surveys, we generated prevalence ratios as measures of the effect of selected potential determinants of HCV seropositivity. We used univariable and multivariable modified Poisson regression models with generalized estimating equations, specifying exchangeable correlation structures to allow robust variance estimation and take clustering into account [38-40]. The following predictors of HCV seropositivity were selected a priori based on their potential association with HCV: year of data collection, lifetime history of IDU, HIV seropositivity, history of syphilis (defined as a reactive serological test), age (categorical), birth outside of Canada, annual income ≥30,000 Canadian dollars (CAD), education level higher than high school, sexual orientation other than gay/homosexual, self-identification as Indigenous, first language other than French or English, and specific sexual practices reported in the past P6M: transactional sex (defined as having given/received money, drugs, or other goods or services in exchange for sex), condomless anal sex (CAS) with a man of serodiscordant/unknown HIV status, >5 male sexual partners, and group sex. We used multiple imputation by chained equations for missing values as 33% of observations had at least one missing value for the variables used in these analyses, principally information regarding (23%) the question related to CAS [41, 42].

We conducted additional analyses to investigate whether the association of sexual practices with HCV seropositivity varied by HIV status, and to quantify the joint association of lifetime IDU and sexual behaviours with the outcome. We assessed the presence of additive and multiplicative interactions between the following terms: i) HIV seropositivity and four sexual risk factors reported in the P6M (i.e., transactional sex, CAS with a man of serodiscordant/unknown HIV status, number of sexual partners, and group sex) and ii) history of IDU and the same four factors. To evaluate multiplicative interaction, we included the product term of each selected pair of factors in the main analysis model separately and examined the estimate and CI of each product term coefficient [43]. Additive interaction was assessed by transforming the parameter estimates to obtain the relative excess risk due to interaction (RERI) using the method developed by Hosmer and Lemeshow [44]. CIs for the RERIs were computed following Zou [45]. All analyses were conducted using R 3.5.3 [46], with the *RDS* [47], *geepack* [48], *mice* [49], and *dplyr* [50] packages.

### Ethical considerations

For each survey, written informed consent was obtained from all participants. A $10 CAD compensation was offered to participants of the two Argus surveys. In Engage, both participation ($50 CAD) and recruitment of peers ($15 CAD per recruit) were incentivized. The research ethics board of the *Research Institute of the McGill University Health Center* approved all three surveys.

## Results

### Characteristics of participants (pre-standardization)

A total of 4,139 MSM were included in our analyses (1,795 in 2005, 1,258 in 2009, and 1,086 in 2018) (Figure 1). Before standardization, the compositions of the three samples differed. The median age was higher in 2005 and 2009 than in 2018 (Table 1). The proportion of men born outside of Canada was twice as low in the 2005 and 2009 samples as in the 2018 sample. The 2009 sample further contrasted with the others regarding social factors, with participants reporting a higher income, a lower level of education, and being more likely to identify as gay/homosexual. The proportion of participants living with HIV increased from 13% (95%CI: 11-14%) in 2005, to 15% (95%CI: 13-17%) in 2009, and 18% (95%CI: 16-21%) in 2018. We observed no clear trend in the percentage of participants with a history of IDU: 7% in 2005 (95%CI: 6-8%), 12% (95%CI: 10-14%) in 2009, and 10% (95%CI: 8-12%) in 2018. However, the proportions of MSM who had a reactive serological test for syphilis, reported recent CAS with a man of serodiscordant/unknown HIV status, and reported >5 male sexual partners in the P6M increased over time. Conversely, recent engagement in transactional sex decreased over the study period. Across the three surveys, 61% of participants who reported transactional sex gave, 62% received, and 23% both gave and received money, drugs, or other goods and services in exchange for sex. The proportion of MSM who engaged in group sex in the P6M increased from 2005-2009, and then decreased in 2018.

**Table 1.** Description of participants included in three cross-sectional bio-behavioral surveys of men who have sex with men conducted in Montréal, Québec (Canada; 2005-2018; before any imputation or standardization).

| Characteristic | 2005<br>(N = 1,795)<br>n (%) | 2009<br>(N = 1,258)<br>n (%) | 2018<br>(N = 1,086)<br>n (%) |
| --- | --- | --- | --- |
| HCV seropositivity | 95 (5%) | 49 (4%) | 58 (5%) |
| HIV seropositivity | 224 (13%) | 193 (15%) | 200 (18%) |
| Reactive syphilis serology | 93 (5%) | 116 (9%) | 194 (18%) |
| Age | 38 (28-47)<sup>†</sup> | 40 (30-47)<sup>†</sup> | 34 (28-49)<sup>†</sup> |
| Income ≥30,000 CAD in the past year | 891 (50%) | 763 (61%) | 461 (42%) |
| Education level higher than high school | 1,217 (68%) | 537 (43%) | 806 (74%) |
| Sexual orientation other than gay/homosexual | 336 (19%) | 107 (9%) | 205 (19%) |
| Born outside of Canada | 244 (14%) | 194 (15%) | 348 (32%) |
| **Self-reported ethnicity or family background** |  |  |  |
| Indigenous | 33 (2%)<sup>‡</sup> | 16 (1%) | 9 (1%) |
| English Canadian | 146 (8%)<sup>‡</sup> | 108 (9%) | 103 (9%) |
| French Canadian | 1,219 (68%)<sup>‡</sup> | 865 (69%) | 537 (49%) |
| European | 105 (6%)<sup>‡</sup> | 119 (9%) | 157 (14%) |
| Asian | 31 (2%)<sup>‡</sup> | 14 (1%) | 40 (4%) |
| Arab or North-African | 28 (2%)<sup>‡</sup> | 20 (2%) | 44 (4%) |
| Sub-Saharan African | 3 (<1%)<sup>‡</sup> | 3 (<1%) | 9 (1%) |
| Latin, South or Central American | 60 (3%)<sup>‡</sup> | 29 (2%) | 92 (8%) |
| Caribbean | 20 (1%)<sup>‡</sup> | 7 (1%) | 9 (1%) |
| Oceanian (e.g., Australian, Pacific Islander) | 2 (<1%)<sup>‡</sup> | 0 | 1 (<1%) |
| Other | 84 (5%)<sup>‡</sup> | 61 (5%) | 74 (7%) |
| **First language other than French or English** | 125 (7%) | 101 (8%) | 74 (7%) |
| **Transactional sex in the P6M<sup>§</sup>** | 329 (18%)<sup>‡</sup> | 187 (15%)<sup>‡</sup> | 108 (10%) |
| **CAS with a man of serodiscordant/unknown HIV status in the P6M<sup>¶</sup>** | 200 (11%)<sup>‡</sup> | 251 (20%)<sup>‡</sup> | 395 (36%)<sup>‡</sup> |
| **> 5 male sexual partners in the P6M** | 664 (37%) | 569 (45%) | 533 (51%) |
| **Group sex in the P6M** | 479 (27%) | 407 (32%)<sup>‡</sup> | 252 (23%) |
| **History of injection drug use<sup>\*</sup>** | 128 (7%)<sup>‡</sup> | 147 (12%) | 110 (10%) |

CAD: Canadian dollars, CAS: condomless anal sex, P6M: past six months.
<sup>†</sup> Median (interquartile range).
<sup>‡</sup> More than 2% of observations were missing for this variable, in this dataset.
<sup>§</sup> Transactional sex was defined as having given/received money, drugs, or other goods or services in exchange for sex in the past 6 months.
<sup>¶</sup> Male sexual partners included both oral and anal sexual partners.
<sup>\*</sup> Lifetime injection of any non-prescribed drug was considered as a previous experience of injection drug use.

**Figure 1.**
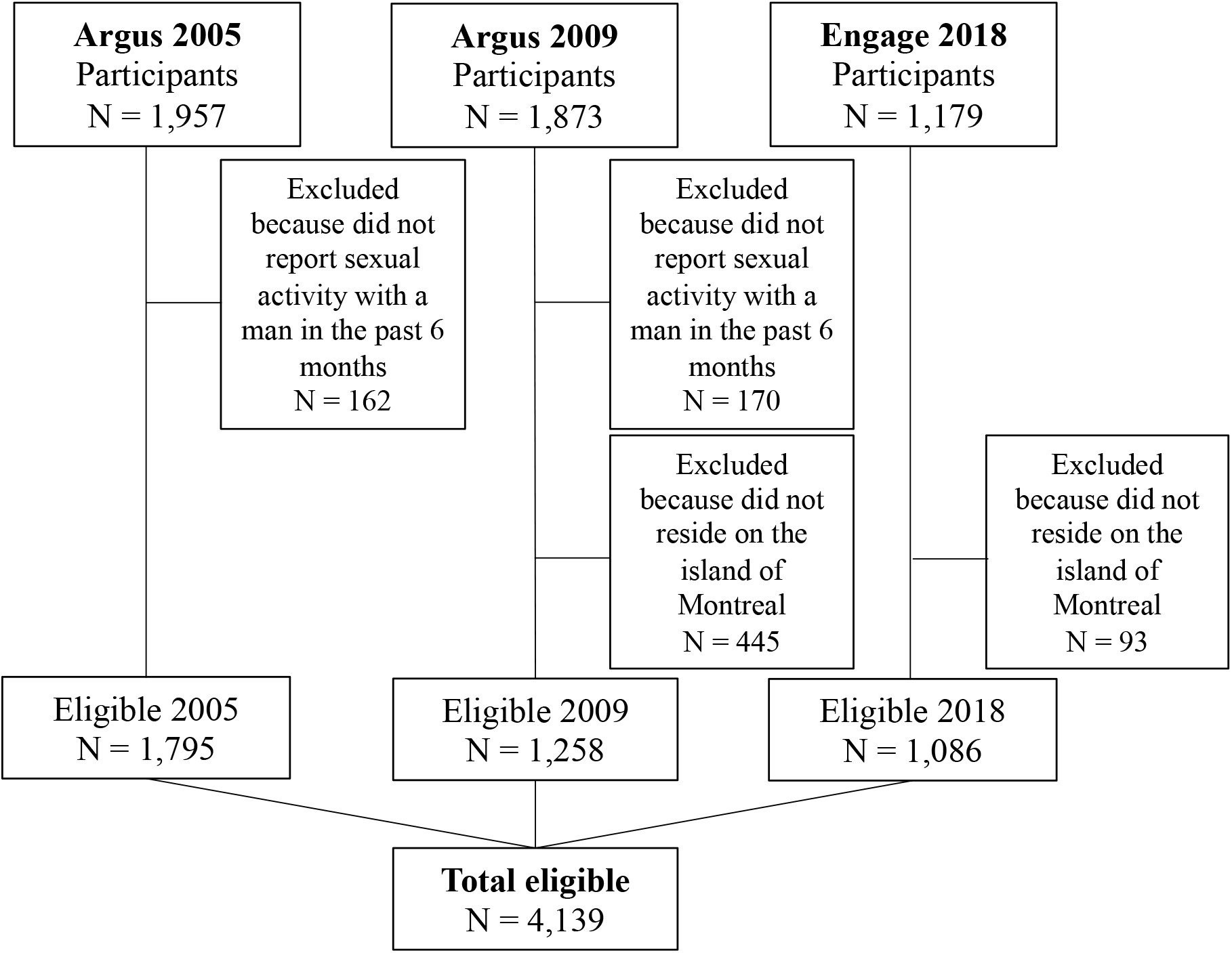
Study eligibility flowchart. Individuals missing values on any eligibility criterion were excluded from our analyses.

### Temporal trends in HCV seroprevalence (post-standardization)

Standardized HCV seroprevalence among all MSM remained stable and ranged from 7% (95%CI: 3-10%) in 2005, to 8% (95%CI: 1-9%) in 2009, and 8% (95%CI: 4-11%) in 2018 (Figure 2, Additional file 2).

**Figure 2.**
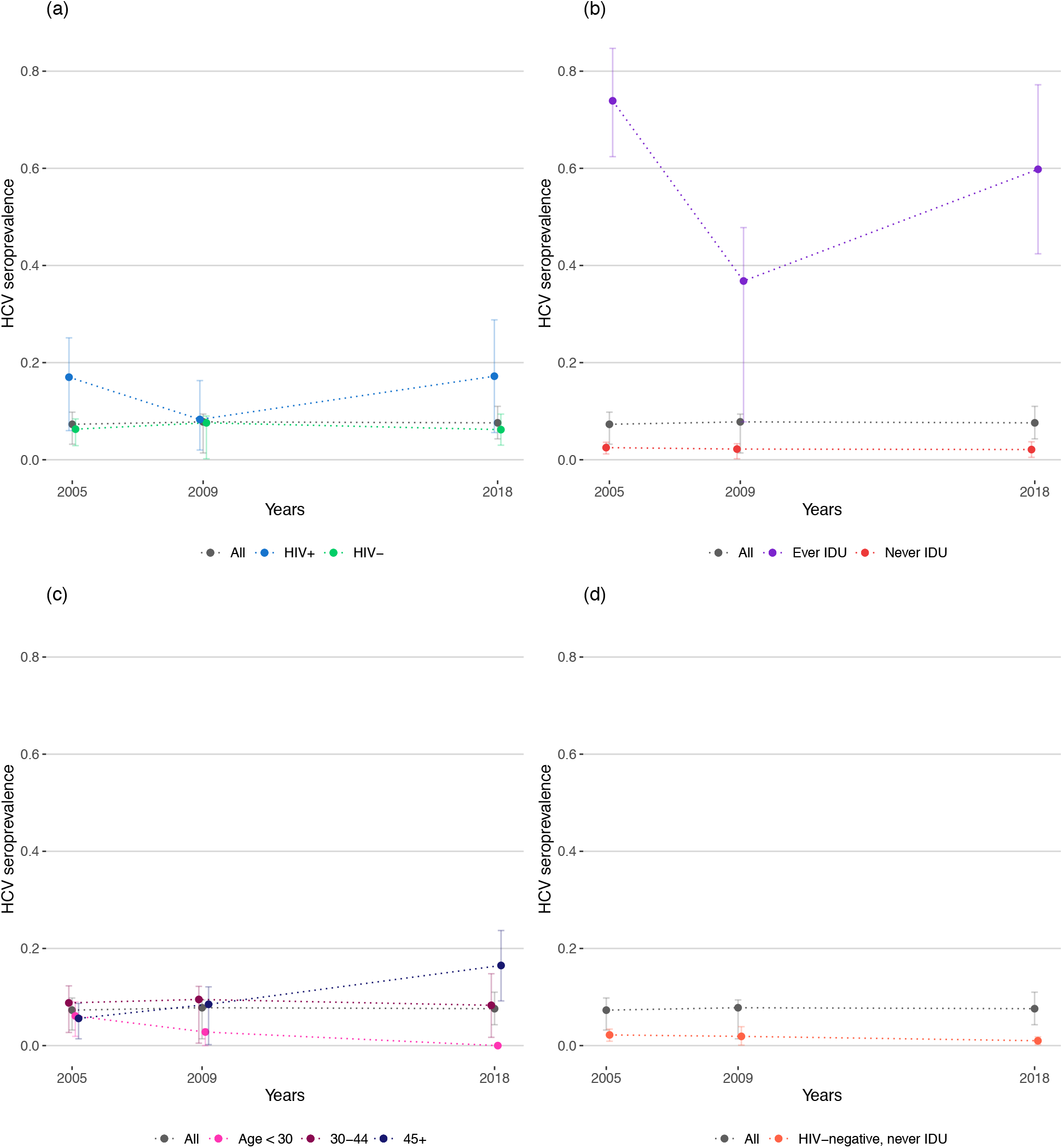
HCV seroprevalence estimates among gay, bisexual, and other men who have sex with men (MSM) overall, stratified by (a) HIV status, (b) lifetime history of injecting drug use (IDU), and (c) age group, and among (d) HIV-negative MSM without a history of IDU in 2005, 2009, and 2018 in Montréal, Québéc (Canada).

Stratification by HIV and IDU status did not reveal consistent temporal trends in HCV seroprevalence among these subgroups. We observed a drop in estimates for MSM living with HIV and MSM with a history of IDU in 2009 and obtained relatively wide CIs among these two groups due to smaller numbers of observations. The highest HCV seroprevalence point estimates were obtained among participants with a history of IDU, with 74% (95%CI: 62-85%) in 2005, 37% (95%CI: 8-48%) in 2009, and 60% (95%CI: 42-77%) in 2019. In contrast, HCV seroprevalence among MSM without a history of IDU was estimated at 3% (95%CI: 1-4%) in 2005, 2% (95%CI: 0-3%) in 2009, and 2% (95%CI: 1-4%) in 2018. HCV seropositivity was also far more common among MSM living with HIV at 17% (95%CI: 6-25%) in 2005, 8% (95%CI: 2-16%) in 2009, and 17% (95%CI: 6-29%) in 2018, than among HIV-negative MSM at 6% (95%CI: 3-8%) in 2005, 8% (95%CI: 0-9%) in 2009, and 6% (95%CI: 3-9%) in 2018. Finally, HCV seroprevalence remained relatively stable among HIV-negative MSM without a history of IDU at 2% (95%CI: 1-3%) in 2005, 2% (95%CI: 0-4%) in 2009, and 1% (95%CI: 0-2%) in 2018.

When stratifying by age group, we observed a decrease in HCV seroprevalence among MSM <30 years old over time, from 6% (95%CI: 2%-9%) in 2005, to 3% (95%CI: 0-3%) in 2009, and 0% (95%CI: 0-1%) in 2018. We noted no clear temporal trend in HCV seroprevalence among MSM aged 30-44 years (9%; 95%CI: 3-12% in 2005, 10%; 95%CI: 1-12% in 2009, and 8%; 95%CI: 2-15% in 2018). Among MSM aged ≥45 years, HCV seroprevalence increased from 6% (95%CI: 1-9%) in 2005 to 9% (95%CI: 0-12%) in 2009 and to 17% (95%CI: 9-24%) in 2018.

### Factors associated with HCV seropositivity

Lifetime history of IDU was the strongest predictor of HCV seropositivity in our population (Table 2). Increased age, recent transactional sex, HIV seropositivity, and sexual orientation other than gay/homosexual were also associated with increased HCV seroprevalence. Our results suggest an association between socio-economic factors and the outcome: MSM with higher income and education level were less likely to test HCV-seropositive. Being born outside of Canada was also associated with lower HCV seroprevalence. Few participants identified as Indigenous (1-2% across surveys), leading to wide uncertainty around the aPR for this population group. We found no association between recent sexual behaviours studied and HCV seropositivity. Finally, first language other than French or English was not associated with a variation in the outcome.

**Table 2.** HCV seroprevalence ratios obtained by pooling three cross-sectional surveys of men who have sex with men conducted in Montréal, Québec (Canada) (2005, 2009, 2018).

| **Covariates** | **Univariable prevalence ratio**<br>**(95% confidence interval)** | **Multivariable prevalence ratio**<br>**(95% confidence interval)** |
| --- | --- | --- |
| **History of IDU** | 18.2 (12.2-27.2) | 8.0 (5.5-11.5) |
| **Age** |  |  |
| <30 | 1.00 | 1.00 |
| 30-44 | 2.1 (1.4-3.2) | 2.2 (1.5-3.3) |
| ≥45 | 1.9 (0.9-4.1) | 2.5 (1.6-3.9) |
| **Transactional sex (P6M)** | 4.6 (3.0-7.1) | 2.1 (1.7-2.5) |
| **HIV seropositivity** | 2.9 (1.8-4.8) | 1.7 (1.3-2.3) |
| **Sexual orientation other than**<br>**gay/homosexual** | 5.4 (3.2-9.2) | 1.6 (1.3-2.1) |
| **Income ≥30,000 CAD in the past**<br>**year** | 0.3 (0.2-0.4) | 0.5 (0.4-0.7) |
| **Born outside of Canada** | 0.2 (0.0-0.6) | 0.5 (0.3-0.9) |
| **Year of data collection** |  |  |
| 2005 | 1.0 | 1.0 |
| 2009 | 0.4 (0.2-0.8) | 0.5 (0.3-0.8) |
| 2018 | 0.9 (0.3-2.4) | 1.0 (0.7-1.4) |
| **Education level higher than high**<br>**school** | 0.2 (0.1-0.3) | 0.4 (0.3-0.6) |
| **Self-identified Indigenous**<br>**ethnicity or family background** | 1.1 (0.1-23.7) | 1.4 (0.4-4.5) |
| **Reactive syphilis serology** | 0.9 (0.4-1.9) | 1.1 (0.7-1.6) |
| **1<sup>st</sup> language other than French**<br>**or English** | 0.4 (0.1-1.0) | 1.0 (0.5-2.0) |
| **> 5 male sexual partners (P6M)** | 0.8 (0.5-1.4) | 1.0 (0.7-1.3) |
| **Group sex in the P6M** | 0.8 (0.5-1.2) | 0.8 (0.6-1.2) |
| **CAS with a man of**<br>**serodiscordant/ unknown HIV**<br>**status (P6M)** | 0.7 (0.3-1.6) | 0.7 (0.5-1.1) |

CAD: Canadian dollars; CAS: condomless anal sex; IDU: injecting drug use; P6M: past 6 months.

### Sensitivity analyses

We detected negative additive and multiplicative interactions between HIV seropositivity and transactional sex in the P6M (Table 3). The presence of negative additive and multiplicative interactions means that the association with HCV seropositivity when both factors are present is smaller than, respectively, the sum and the product, of the individual associations of the two factors with the outcome. We also identified negative additive interactions between history of IDU and each one of the sexual risk factors studied, separately, and a positive multiplicative interaction between history of IDU and group sex in the P6M. A positive multiplicative interaction indicates that the association when both factors are present is stronger than the product of the individual associations.

**Table 3.** Adjusted prevalence ratios of multiplicative interaction terms and relative excess risk due to interaction of selected potential risk factors for HCV seropositivity pooling three cross-sectional surveys of men who have sex with men in Montréal, Québéc (Canada; 2005, 2009, 2018).

| Interaction studied | Exponentiated coefficient of the multiplicative interaction term (95% confidence interval) | Relative excess risk due to interaction (95% confidence interval) |
| --- | --- | --- |
| HIV infection; CAS with a man of serodiscordant/ unknown HIV status (P6M) | 1.0 (0.4; 2.3) | -0.5 (-1.5; 0.9) |
| HIV infection; >5 male sexual partners (P6M) | 0.9 (0.5; 1.6) | -0.8 (-1.7; 0.1) |
| HIV infection; transactional sex (P6M) | 0.6 (0.4; 0.9) | -3.3 (-4.6; -2.3) |
| HIV infection; group sex (P6M) | 0.9 (0.5; 1.7) | -0.8 (-1.7; 0.2) |
| History of IDU; CAS with a man of serodiscordant/ unknown HIV status (P6M) | 1.0 (0.5; 1.9) | -6.7 (-10.2; -4.1) |
| History of IDU; >5 male sexual partners (P6M) | 1.1 (0.7; 1.8) | -6.5 (-10.6; -3.8) |
| History of IDU; transactional sex (P6M) | 0.9 (0.6; 1.6) | -8.9 (-13.9; -5.6) |
| **History of IDU; group sex (P6M)** | **2.0 (1.1; 3.7)** | **-4.6 (-7.9; -1.7)** |

CAS: condomless anal sex; IDU: lifetime injecting drug use; P6M: past 6 months.
These interaction terms were included in the multivariable model described in table 4, separately.
All the variables used in the interaction terms are binary (coded 0,1).

## Discussion

Standardizing and pooling three large cross-sectional surveys of MSM from 2005-2018, our results suggest that HCV seroprevalence has remained stable at approximately 8% in Montréal. Overall, the point estimates for HCV seroprevalence among all MSM are higher than those recently estimated in other North American settings: 5% in Vancouver [15], 4% in San Francisco, California [51], and 3% in New-York City [52]. Pre-standardization estimates of HCV seroprevalence among all MSM were also stable over the study period at approximately 5%, contrasting with the increase observed in the prevalence of HIV and biological markers of syphilis.

Despite the overall stability in HCV seroprevalence through time, we observed diverging trends among age groups. Because Canadian public health authorities do not systematically collect HCV incidence data, monitoring changes in HCV seroprevalence among young adults has been proposed as an approach to gauge trends in HCV incidence in the general population [8]. The decrease observed in HCV seroprevalence among participants <30 years old may therefore indicate a decline in new HCV infections among MSM from 2005-2018. In contrast, the increase in HCV seroprevalence among MSM aged ≥45 years may reflect birth cohort effects (i.e., males born between 1945-1975 [2]), temporal changes in injecting and/or sexual behaviours among older MSM, or changes in inter-generational transmission of HCV over time.

HCV seroprevalence was particularly high among MSM living with HIV, and even more so among MSM with a history of IDU. The drop in seroprevalence observed in 2009 among these groups could reflect temporal changes in injecting behaviours (e.g., increased number of new injectors that have never been exposed to HCV). However, we cannot rule out that these results could be due to remaining differences in the composition of the samples post-standardization.

HCV seroprevalence among HIV-negative MSM in Montréal ranged from 6-8% between 2005-2018, on the high end of estimates from Canada [16], the United States [52], and Europe [13]. The seroprevalence estimated among MSM without a history of IDU (2-3%) was also twice as high as that estimated in Vancouver for the same group [15]. These numbers are greater than the 1% HCV seroprevalence estimated for the general Canadian population [2]. Yet, the seroprevalence observed among HIV-negative MSM without a history of IDU was stable in the range of 1-2%, comparable to that among the general population. The relatively high HCV seroprevalence observed among all MSM may therefore reflect frequent exposure to the virus among subgroups with particular vulnerabilities or risk behaviours such as MSM living with HIV, and MSM with a history of IDU.

Reporting a history of IDU was the strongest predictor of HCV seropositivity among Montréal MSM over the study period. This is particularly concerning given that the proportion of MSM who reported lifetime IDU in Engage (6%, [32]) is twelve times higher than that reported among the general population in Montréal (0.5%, [53]). Aside from recent transactional sex, which encompasses having given/received drugs (or money, goods, or services) in exchange for sex and has been previously identified as an important risk factor for HCV among MSM in Canada [15], we found no evidence of an association between sexual behaviours and HCV seroprevalence. These results are consistent with those obtained in Toronto [16], and with a study conducted in 2001 that did not find evidence of sexual transmission of HCV among a cohort of Montréal MSM [28]. However, they contrast with emerging evidence from Vancouver [20] and with studies conducted in Europe [9] and the United States [17], where sexual risk factors were strongly associated with new HCV infections among MSM. One potentially important risk factor that we could not investigate is chemsex, a form of sexualized drug use that has been previously associated with a higher risk of HCV infection [54, 55]. Nevertheless, the positive multiplicative interaction observed between group sex and IDU could partly capture the association of chemsex practices with HCV seroprevalence, although we could not measure the concomitance of injecting and sexual behaviours.

Altogether, higher socio-economic status (i.e., income and education) was associated with lower HCV seroprevalence. Our data did not allow us to specifically examine the relationship between birth in HCV endemic countries and HCV seropositivity. In this regard, interpreting the negative association between birth outside of Canada and HCV seroprevalence is challenging. Having a first language other than French or English, another factor used as a proxy for country of origin, was not associated with the outcome. Reporting a sexual orientation other than gay/homosexual (e.g. bisexual, heterosexual, queer) was positively associated with HCV seropositivity. In line with previous work, this finding suggests that sexual orientation and behaviours should be examined in conjunction in order to fully understand how sexually transmitted and blood-borne infections are distributed among MSM populations [56].

Our results should be interpreted considering the study’s main limitations. First, the use of cross-sectional data renders the interpretation of observed associations challenging due to a temporality bias. However, pooling the available cross-sectional data allowed us to produce robust, comparable estimates of HCV exposure across time, and to investigate the plausibility of different modes of transmission of the virus among these groups. Second, we could not investigate the relationship between lifelong sexual practices and HCV seropositivity due to data limitations. Yet, we were able to quantify the association of numerous recent sexual behaviours with HCV exposure among Montréal MSM, adjusting for important factors such as age.

Strengths of this study include its large sample size, resulting from the pooling of three comparable survey instruments. No sampling framework is available for MSM populations, and the Engage survey used an RDS design to constitute a diverse sample of MSM that enabled us to adjust for potential sampling biases in the Argus time-location surveys. Additionally, recent epidemiological data on HCV exposure and determinants among MSM living in Montréal is lacking, and very few population-based studies have examined HCV prevalence trends among MSM subgroups globally. As such, our work provides a unique opportunity to fill these research gaps.

## Conclusions

HCV seroprevalence remained high among Montréal MSM between 2005-2018, likely reflecting frequent exposure to the virus among MSM living with HIV and MSM with a history of IDU. Unlike other settings where HCV infection was strongly associated with sexual risk factors among MSM subgroups, history of IDU was the preeminent risk factor for HCV seropositivity among Montréal MSM. Understanding the intersection of IDU contexts, practices, and populations is essential to prevent HCV transmission among these overlapping populations.

## Data Availability

The data referred to in the manuscript are not publicly available.

## Authors’ contributions

C.L.D., J.C., M.K., and M.M.-G. conceived and designed the study. J.C. and G.L. were involved in the study design and data collection of the Argus 1 and 2 surveys. J.C., G.L., D.G., and N.L. were involved in the study design and data collection of the Engage study. C.L.D. administered and processed the different databases. C.L.D. performed the statistical analyses with inputs from M.K. and M.M.-G. All authors contributed to results interpretation. C.L.D. drafted the manuscript and all authors critically reviewed it for important intellectual content. All authors approved the final version.

## Acknowledgements

The principal investigators of Argus are Michel Alary, Chris Archibald, Joseph Cox, Louis-Robert Frigault, Marc-André Gadoury, Gilles Lambert, René Lavoie, Robert Remis, Paul Sandstrom, Cécile Tremblay, François Tremblay, and Jon Vincelette. We would like to thank all the men who agreed to participate in the Argus surveys, as well as the owners and managers of the establishments where the participants were recruited. We would also like to thank the gay community organizations in Montréal who supported this research. The principal investigators of Engage are Joseph Cox, Daniel Grace, Trevor Hart, Jody Jollimore, Nathan Lachowsky, Gilles Lambert, and David Moore. We would like to thank all the Engage participants, the community engagement committee members, and the affiliated community agencies.

## Additional files

Additional file 1: Eligibility criteria for participating in the Argus 1 (2005), Argus 2 (2009), and Engage (2018) studies conducted in Montréal, Québec (Canada).

| **Study** | **Eligibility criteria** |
| --- | --- |
| **Argus 1 (2005)** | - Gender-identify as a man<br>- Have ever had sex with a man<br>- Be 18 years of age or older<br>- Speak French or English<br>- Reside on the island of Montréal<br>- Be able to provide informed consent |
| **Argus 2 (2009)** | - Gender-identify as a man<br>- Have ever had sex with a man<br>- Be 18 years of age or older<br>- Speak French or English<br>- Reside in the province of Quebec<br>- Be able to provide informed consent |
| **Engage (2018)** | - Gender-identify as a man<br>- Report sexual activity with a man in the past 6 months<br>- Be 16 years of age or older<br>- Be able to complete a computer-based questionnaire in French or English<br>- Reside in the greater Montréal region<br>- Be able to provide informed consent<br>- Receive a voucher for participation in the study, or be purposively invited to be an initial “seed” |

Additional file 2: Standardized HCV seroprevalence among all men who have sex with men (MSM) and stratified by HIV status, by injection drug use status, and by age group, in 2005, 2009, and 2018 in Montréal, Québec (Canada).

| **Group** | **Standardized HCV seroprevalence (95% confidence interval)** |  |  |
| --- | --- | --- | --- |
|  | **2005** | **2009** | **2018** |
| **All MSM** | 7% (3-10) | 8% (1-9) | 8% (4-11) |
| **MSM living with HIV** | 17% (6-25) | 8% (2-16) | 17% (6-29) |
| **HIV-negative MSM** | 6% (3-8) | 8% (0-9) | 6% (3-9) |
| **MSM with a history of IDU** | 74% (62-85) | 37% (8-48) | 60% (42-77) |
| **MSM without a history IDU** | 3% (1-4) | 2% (0-3) | 2% (1-4) |
| **MSM aged <30 years old** | 6% (2-9) | 3% (0-3) | 0% (0-1) |
| **MSM aged 30-44 years old** | 9% (3-12) | 10% (1-12) | 8% (2-15) |
| **MSM aged ≥45 years old** | 6% (1-9) | 9% (0-12) | 17% (9-24) |

| Group | Standardized HCV seroprevalence (95% confidence interval) |  |  |
| --- | --- | --- | --- |
|  | 2005 | 2009 | 2018 |
| **HIV-negative MSM without a history of IDU** | 2% (1-3) | 2% (0-4) | 1% (0-2) |

IDU: injection drug use; MSM: men who have sex with men.

## List of abbreviations

aPR: adjusted prevalence ratio
CAD: Canadian dollars
CAS: condomless anal sex
CI: confidence interval
HCV: hepatitis C virus
HIV: human immunodeficiency virus
MSM: gay, bisexual, and other men who have sex with men
P6M: past six months
RDS: respondent-driven sampling
RERI: relative excess risk due to interaction
RNA: ribonucleic acid
WHO: World Health Organization

